# Molecular Detection of *bla* OXA-48 Gene Encoding Carbapenem Resistance Pseudomonas aeruginosa Clinical Isolates from Khartoum State Hospitals, Sudan

**DOI:** 10.1101/2020.06.22.20137034

**Authors:** Doha Omer Ali, Mohamed M.A. Nagla

## Abstract

Carbapenem resistance in *Pseudomonas*.*aeruginosa* is particularly worrisome because this class of β-lactam represents the last therapeutic resource for control of bacterial infection.So this study aimed to detect the frequency of *bla* OXA-48 resistance gene among *Pseudomonas aeruginosa* clinical isolates during the period from November 2018 to November 2019.

Hundred *Pseudomonas aeruginosa* clinical isolates, 81 carbapenems (imipenem meropenem) resistant and 19 carbapenems sensitive were collected from Omdurman Teaching Hospital, Fedail Hospital and Soba Teaching Hospital in Khartoum State-Sudan. All isolates were re-identified using conventional bacteriological techniques, their susceptibility to carbapenems were tested using Kirby-Bauer method for confirmation and investigated for the presence of the *bla* OXA-48 gene using conventional PCR technique.

60 (60.0%) out of 100 *Pseudomonas aeruginosa* clinical isolates were positive for *blaOXA*-48 gene. Out of 81 carbapenem resistant isolates 54(66.7%) were positive for *bla* OXA-48 gene, while among the (19) carbapenem sensitive isolates 6 (31.6%) were positive for blaOXA-48 gene. There was statistically significant association between carbapenem resistant isolates and the presence of *bla*OXA-48 gene (*P-value* = 0.006).

Wound swabs were the predominant clinical samples detected harboring *bla* OXA-48 gene both among the sensitive 5 (83.3%) and carbapenem resistant isolates 29(53.7) (P.value> 0.05).

Our findings revealed high frequency of *bla* OXA-48 among carbapenem resistant isolates so identification of *bla* OXA-48 producing strains and taking efforts to reduce the rate of transferring these gene between the different strains is essential for optimization of therapy and improves of patients outcomes.

## Introduction

*Pseudomonas aeruginosa* (*P*.*aeruginosa*) is gram negative bacterium.[1]. It is a lethal opportunistic pathogen that is a leading cause of morbidity and mortality in immunocompromised individuals [2]. It rarely found in the microbiota of healthy humans [3].The colonization rate by by *P*.*aeruginosa* significantly increases (reaching up to 80%) in patients with chronic illnesses (e.g cystic fibrosis, sever burns) or extensive exposure to health care facilities involving interruption of protective barriers [4, 5]. It has an intrinsic resistance to wide range of antimicrobial agents and a high capacity to attain resistance mutations and mobile genetic elements [6]. Elimination of *P. aeruginosa* has become increasingly difficult due to its remarkable capacity to resist antibiotics [2].

Carbapenem resistance in *P*.*aeruginosa* is particularly worrisome because this class of β-lactam represents the last therapeutic resource for control of bacterial infection [7]. Although porins loss and efflux pumps may contribute to the phenotype of carbapenem resistance in *P*.*aeruginosa*, Yet the most relevant resistance mechanism is the production of carbapenem –hydrolyzing enzymes (carbapenemase)[7].

Carbapenemase producing carbapenem resistance enterobacteracea (CP-CRE) that directly break down carbapenem have been classified to various types such as class A β-lactamase (NMC, IMI, SMS, MKPC, GES), class B β-lactamase (IMP, VIM, SPM, NDM) and class D β-lactamase (OXA-48) [8]. In recent years, the emergence of diverse carbapenemase in members of the family Enterobacteriaceae has become a major challenge for health care system [9].

BlaOXA-48 gene is a class D β-lactamase that possesses the ability to hydrolyze carbapenems as well as Cloxacillin and Penicillin [10]. Studies done by Carrër, et al., (2010) indicated that the blaOXA-48 gene has been found in plasmid and located between two identical insertion sequences, IS999, forming the composite transposon Tn 1999s [11]. The genetic environment of blaOXA-48 was investigated by PCR with primers specific for insertipon sequence IS1999 and for the 5′ part of blaOXA-48 [11]. Recently, Nordmann et al described a new inhibitor-based biochemical assay for carbapenemase detection [12]. But detection of OXA-48 and related enzymes remains problematic, as no specific inhibitor is available. Temocillin resistance was suggested as an indicator of OXA-48 production but not for OXA-48 confirmation [13-15].

The first case of resistance mediated by OXA-48 was identified in 2001 from multi-drug resistance *Klebsiella pneumoniae* in Istanbul Turkey [16]. Several cases of OXA-48 producing organisms like *Pseudomonas, E*.*coli* and Cetrobacter were reported in patients from or have relation with Turkey [17].Several isolates producing OXA-48 have been involved as a source of nosocomial outbreaks [18, 19].Outbreak of OXA-48 was reported worldwide, disseminating from Middle Eastern countries and North Africa. Sporadic cases have been reported in Lebanon, Oman, Saudi Arabia and Kuwait. In Africa mostly in Northern African countries of Egypt, Tunisia, Libya and Morocco [20].And little is known about this problem in Sudan, as a rapid and reliable detection of carbapenemases is desirable in order to limit the spread of these enzymes Thus the present study aimed to investigate the frequency of blaOXA-48 resistant gene among carbapenem resistance *P. aeruginosa*.

## Materials and Methods

### Design and setting

A descriptive - cross sectional study was conducted in different Khartoum State hospitals, Sudan (Omdurman Teaching Hospital, Fedail Hospital and Soba Teaching Hospital) during the period from November 2018 to November 2019. A total of 100 *Pseudomonas aeruginosa* clinical isolates of known carbapenem susceptibility from different types of clinical samples (wound swabs, urine, ear swabs, sputum and high vaginal swabs) were included in this study. They represented 81 carbapenems (imipenem meropenem) resistant and 19 carbapenems sensitive.

*Pseudomonas aeruginosa* clinical isolates of known resistant and sensitive to both routinely used carbapenem (Meropenem and Imipenem) antibiotics were included in this study and any clinical isolates resist or sensitive to only one carbapenem antibiotics was excluded from this study. The approval and informed consent was obtained from Faculty and Department of Medical Microbiology management, Al-Neelain University and informed consent was taken from Hospitals included in this study.

### Phenotypic methods

The fresh cultured colonies of *P*.*aeruginosa* clinical isolates were sub-cultured on nutrient agar and macConkey agar media to ensure purity and optimal growth. Then they were re-identified using routine culture techniques and their susceptibility to carbapenems were tested using Kirby-Bauer method for confirmation.

### DNA extraction

Bacterial DNA was extracted by using boiling method. All bacterial strains were sub cultured on nutrient agar and the overnight pure bacterial growth were used. Three colonies were emulsified in Eppendorf tubes containing 200µl of the distilled water and boiled at 100 °C for 30 minutes then centrifuged at 12000 rmp for 15 minutes. The supernatant containing DNA was transferred to new Eppendorf tubes and stored at -20°C [21]

### PCR amplification procedure

PCR was carried out for all isolates by using specific primer for OXA-48 gene F 5’**GCGTGGTTAAGGATGAACA** 3’ R 5’**CATCAAGTTCAACCCAACCG** 3’ of amplicon size of 177 bp [22]. Amplification was performed at final volume 25 µl (5µl Mater mix, 5µl from DNA, 2µl primer and 13µlD.W).The PCR cycling conditions were as follows: an initial denaturation step of 10 min at 94°C followed by 35 cycles of 30 s at 94°C, 30 s at 55°C and 30s at 72°C,with a final extension of 10 min, (Tc-412,UK) thermo cycler machine was used.

PCR products was separated on 2% agarose gel by electrophoresis to detect specific amplified product by comparing with standard molecular weight marker 100 base pair (DNA ladder) 5µl of PCR product was loaded into the well. Electrophoresis was carried out at 100 Volts, 60 AMP and 30 minutes.

The amplified products of the study samples were visualized by Trans -illuminator. Photographed by a digital camera and transferred to computer data for labeling and storage.

## Results

Hundred (100) *P*.*aeruginosa* clinical isolates were collected from different hospitals in Khartoum State, Sudan during the period from November 2018 to November 2019. They were obtained from different clinical samples. They represented (57) wound swabs, (24) urine, (3) ear swabs, (6) sputum and (10) high vaginal swabs (Table 1).Their identification was confirmed using conventional bacteriological techniques.

**Table 1:**
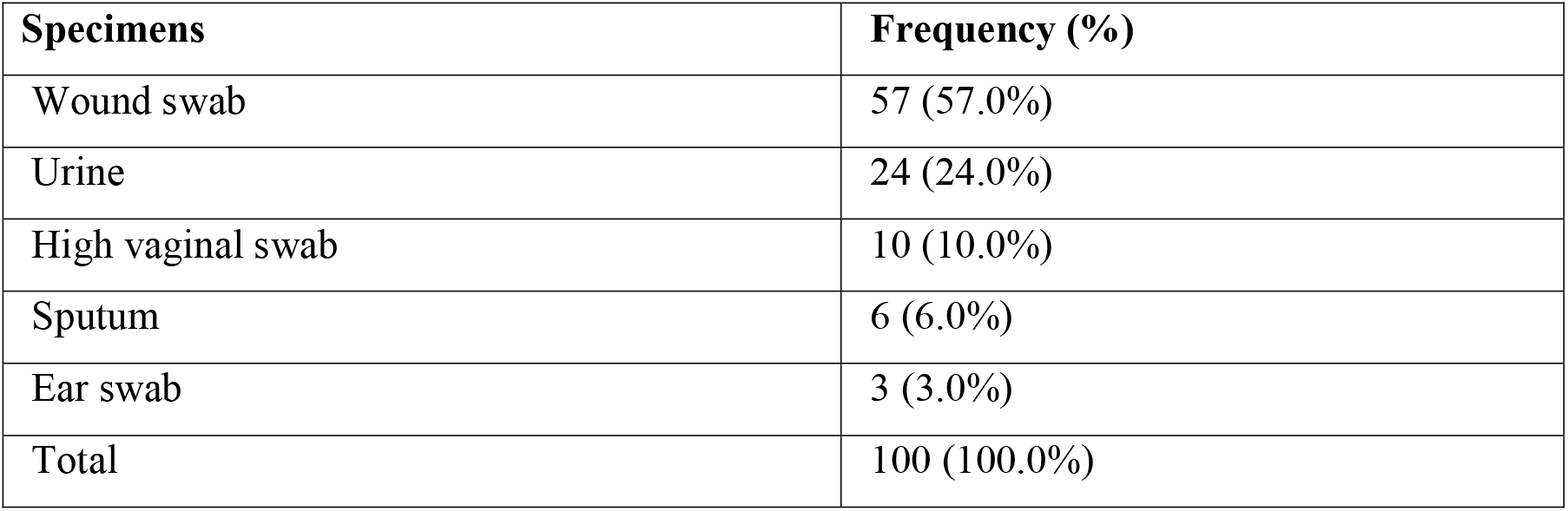
Distribution of *Pseudomonas aeruginosa* clinical isolates according to type of specimens.

Carbapenem (Meropenem and Imipenem) susceptibility testing using disc diffusion method showed that 81(81.0%) of isolates were Meropenem and Imipenem resistant, While 19 (19.0%) were Meropenem and Imipenem sensitive (Figure 1).

**Figure 1:**
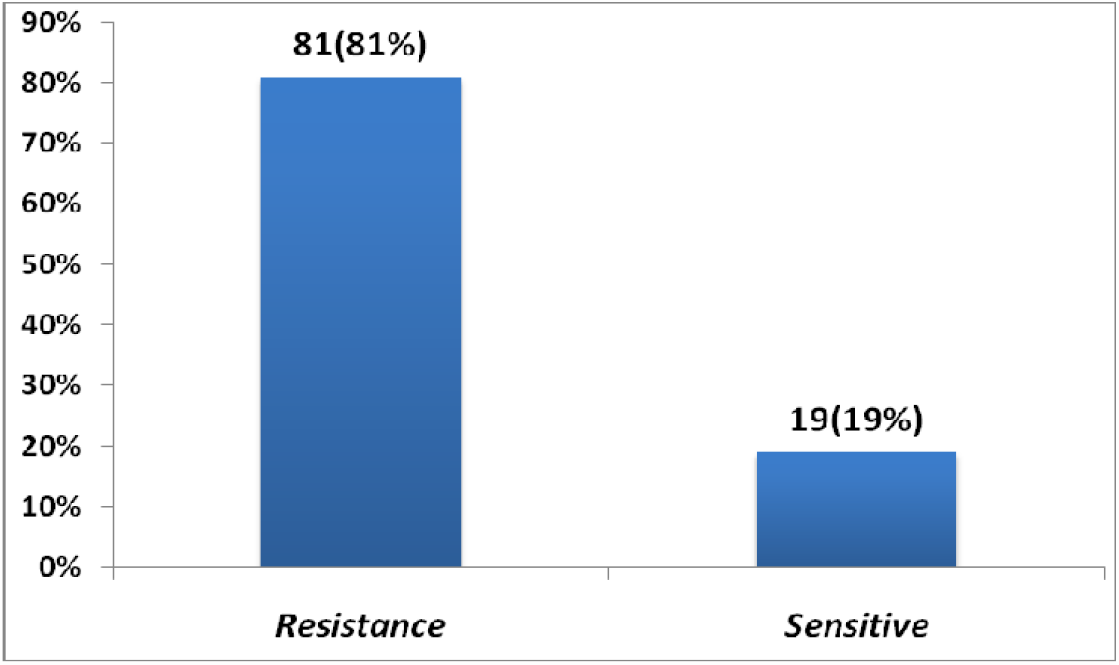
Distribution of carbapenem susceptibility tests among the 100 clinical isolates of *P.aeruginosa*.

Based on PCR result (Figure 2), 60 (60.0%) out of 100 clinical isolates of *Pseudomonas aeruginosa* were positive for *bla*OXA-48 resistance gene (Figure 3).

**Figure 2:**
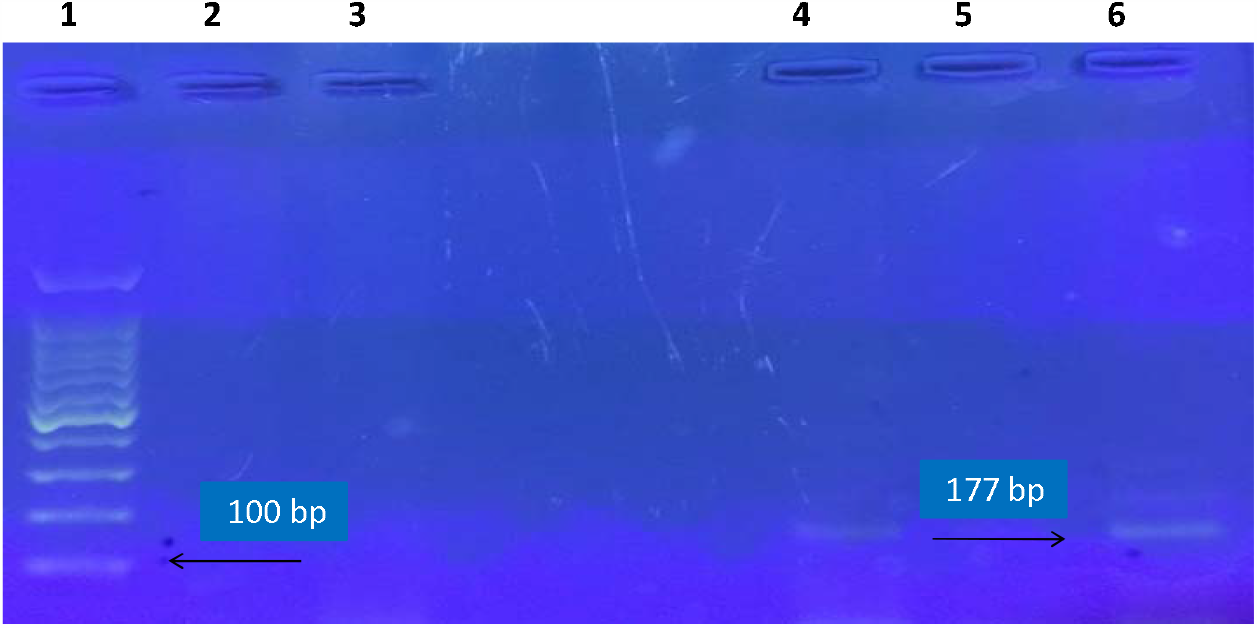
Amplified DNA under ultra violet light machine showing *bla* OXA-48 positive result. Lane 1, DNA leader 100 bp, ;Lanes 2,3, 4, 5 negative samples. Land 6 was a positive sample (typical band size of 177 bp corresponding to the molecularsize of *bla* OXA-48 gene).

**Figure 3:**
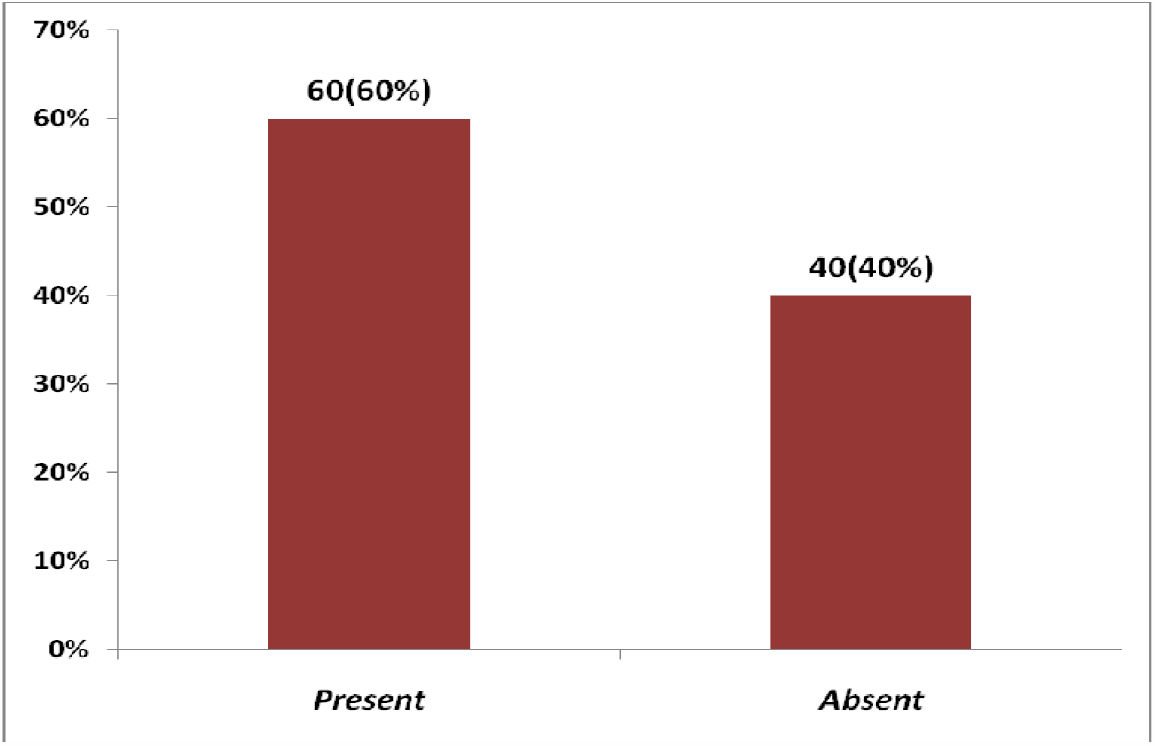
Frequency of blaOXA-48 resistance gene among the 100 *Psaeudomonas aruginosa* clinical isolates.

Out of 81 carbapenem resistant strains 54(66.7%) were positive for *bla* Oxa-48 resistance gene, while among the 19 sensitive strains 6 (31.6%) were positive for blaOXA-48 gene (Table 2). There was a statistically significant association between carbapenem susceptibility and the frequency of *bla*OXA-48 gene with (*P-value* =0.006).

**Table 2:** Frequency of *bla* OXA-48 gene according to carbapenem (Meropenem, Imipenem) susceptibility testing.

| Carbapenem<br>susceptibility test | OXA-48 gene |  | Total |
| --- | --- | --- | --- |
|  | Present | Absent |  |
| Resistance | 54 (66.7%) | 27 (33.3%) | 81 (100.0%) |
| Sensitive | 6 (31.6%) | 13 (68.4%) | 19 (100.0%) |
| Total | 60 (60.0%) | 40 (40.0%) | 100 (100.0%) |
| *P*-value | 0.006 |  |  |

Table (3) showed that the frequency of blaOXA-48 gene among the resistant strains was most common among wound swabs 29 (53.7%) followed by urine 14 (25.9%) (P value > 0.05), similar, it was most frequent among wound swabs 5 (83.3%) followed by urine 1 (16.7%) among the sensitive strains (P value > 0.05) (Table 3-4).There was a statistically insignificant association between the frequency of *bla*OXA-48 gene and the type of specimens.

**Table 3:** Frequency of carbapenemase *bla* OXA-48 gene among the carbapenem resistant strains of *P.aerginosa* according to the type of specimens.

| Specimens | OXA-48 gene |  | Total |
| --- | --- | --- | --- |
|  | Present | Absent |  |
| Wound swab | 29 (53.7%) | 15 (55.6%) | 44 (54.3%) |
| Urine | 14 (25.9%) | 7 (25.9%) | 21 (25.9%) |
| Ear swab | 2 (3.7%) | 1 (3.7%) | 3 (3.7%) |
| Sputum | 5 (9.3%) | 1 (3.7%) | 6 (7.4%) |
| High vaginal swab | 4 (7.4%) | 3 (11.1%) | 7 (8.6%) |
| Total | 54 (100.0%) | 27 (100.0%) | 81 (100.0%) |
| *P-value* | 0.903 |  |  |

## Discussion

Carbapenem resistance mediated by carbapenemases is an important issue in infection control programmers [23]. BlaOXA-48 represents the main enzyme among OXA β-lactamases isolated around the world [24].

In the present study it was found that among the total (100) clinical isolates, the most common isolates were from wound swab specimens, because *Pseudomonas aeruginosa* leading to cause of health care acquired infections most commonly associated with wound infections [25].

In the current study the frequency of *bla* OXA-48 resistance gene was 60% out of 100 *Pseudomonas aeruginosa* clinical isolates. Our result was higher than that study done in Sudan by Mohamed, *et al*., (2018) [26] it was found out of 67 *P. aerugenosa* clinical isolates 22.4% were positive for *bla* OXA-48 resistance gene by real time PCR, This may be due to differences in sample size.

Among (81) carbapenem resistant isolates 54 (66.7%) were positive for *blaOXA*-48 resistance gene (P value < 0.05). It was noted that the absence of *bla* OXA-48 resistance gene among the 27(33.3%) resistant strains may be due to impermeability, which arise via the loss of the Opr D porin, the up-regulation of active efflux pump system present in cytoplasmic membrane of the bacteria and *bla* OXA-48 resistance gene encoded on mobile genetic elements, it has ability to transfer from one bacterium to another [27].Or they may possess other type of carbapenemase enzymes rather than *bla* OXA-48 gene. While among the (19) carbapenem sensitive isolates, 6 (31.6%) were positive for *bla* OXA-48 resistance gene. The presence of resistance gene among the sensitive strains may indicates that there might be a hidden *bla* OXA-48 gene among them which cannot be diagnosed by phenotypic tests as mentioned in study done by Adam *et al*. (2018) [28].

The present study showed that there was a statistically insignificant association between the frequency of *bla*OXA-48 gene and the type of specimens, although there was a high frequency of *bla* OXA-48 resistance gene among wound swabs than other type of specimens. This may be due to the large sample size of wound swabs compared with other specimens, and *P.aeruginosa* was most commonly associated with wound infections.

## Conclusion

In the present work, the significant high frequency of *bla* Oxa-48 resistance gene among the resistance strains may indicates that it is a main source of initiating resistance among these strains, while its presence among carbapenem sensitive isolates suggesting the occurrence of silent gene, that encoded in a movable genetic elements (plasmid), there for, it play a key role in the transfer of horizontal resistance gene from one bacterium to another. Therefore prevention and control programs of carbapenem resistant should be performed to prevent the spread of carbapenemase producers, which includes appropriate use of antimicrobials and facility-level prevention strategies, as recommended by CDC (Centers for Disease Control and prevention).

Wound swabs were the predominant specimens harboring *bla* OXA-48 gene followed by urine samples.

## Data Availability

The data are avialable in the manuscript and note links below

